# ASSESSMENT OF COMPLIANCE TO OCCUPATIONAL HEALTH AND SAFETY SERVICES IN THE ARTISANAL SMALL-SCALE GOLD MINES OF MUMBWA DISTRICT, ZAMBIA

**DOI:** 10.1101/2025.11.24.25340928

**Authors:** Philip Musole, Emmanuel Mulenga, Chritopher Kanema, Patricia Chilaisha, Titus Hamaundu Haakonde, Chisala Deborah Meki, Nosiku Sipilanyambe Munyinda

**Affiliations:** Department of Environmental Health, School of Public Health, University of Zambia, Lusaka, Zambia; Zambia Environmental Management Agency, Lusaka; Department of Occupational Health and safety, Ministry of Labor and Social Services, Zambia

## Abstract

Mercury-dependent gold mining operations expose workers to occupational health and safety hazards due to mercury’s toxicity. This study assessed occupational health and safety (OHS) services provided in mercury-based gold mines in Mumbwa District, Zambia, aiming to evaluate compliance with OHS standards and identify prevalent health hazards faced by miners. A sequential mixed-methods approach was used. Data was collected through questionnaires from 108 conveniently sampled miners, while qualitative data was gathered through in-depth interviews with safety supervisors. Observational checklists were also used in the mines. Workers were exposed to dust and particulate matter (98%), and mercury exposure (62.7%), among other hazards. Compliance with OHS standards was lacking: only 23.8% of mines complied with training standards, and 33.3% with health checks. No mines had onsite medical services. Higher compliance rates were observed for reporting systems (66.7%), first aid services (62%), and fire safety measures (66.7%). Miners in gold amalgamation processes faced mercury exposure, respiratory risks from dust, and environmental contamination. Among workers in mercury-based gold mines, satisfaction was significantly higher among those who received training (OR = 4.82, p = 0.002), had access to First Aid services (OR = 5.88, p = 0.006), and possessed greater experience (OR = 2.75, p = 0.036), while education showed a non-significant trend (OR = 1.92, p = 0.097), and neither gender (OR = 2.25, p = 0.240) nor fire safety measures (OR = 0.96, p = 0.944) were statistically associated with satisfaction. Facilitators to compliance included a safe work environment, regulations and enforcement, and management support, while barriers included financial constraints, lack of training and awareness, and resistance to change. There is an urgent need for enhanced training programs, onsite medical services, and improved worker participation through OHS committees. Comprehensive strategies are needed to improve OHS compliance and protect miners’ health in Mumbwa, District.

**Author Summary:** This study evaluates the level of compliance with occupational health and safety (OHS) protocols among artisanal small-scale gold miners in Mumbwa District, Zambia. Using a mixed-methods design, data were collected from miners to assess adherence to safety standards, availability and use of personal protective equipment, and knowledge of health hazards associated with mining activities. The findings reveal critical gaps in safety compliance, highlighting the need for improved regulatory enforcement, targeted health promotion, and safety training tailored to artisanal miners. This research provides important insights for policymakers and stakeholders to enhance OHS practices, reduce workplace injuries, and promote sustainable mining livelihoods in low-resource settings.

## Introduction

Artisanal and small-scale mining (ASM) is a critical livelihood strategy in sub-Saharan Africa, employing over 20 million workers and supporting 100-150 million dependents (Hilson et al., 2018). The use of mercury in Artisanal and Small-Scale Gold Mining (ASGM) is of increasing international concern. ASGM, while a vital source of income for many rural communities, is the world’s fastest-growing source of mercury contamination (The World Bank, 2013). Globally, 80 to 100 million people depend on ASGM, with an estimated 13 to 15 million artisanal gold miners producing 500 to 800 tons of gold annually while releasing 800 to 1000 tons of mercury. ASGM activities are frequently linked to environmental degradation, poverty, HIV/AIDS, and occupational diseases like pneumoconiosis and silicosis (Basu et al., 2015). Mining operations contaminate the environment by emitting dust particles and chemical emissions into the air, which can cause respiratory impairment (Mwaanga et al., 2019).

Mining is a crucial economic activity in Zambia, but its impact on the environment and workers’ health is concerning. There is a need to examine occupational health and safety systems in relation to safety education among employees and employers. Occupational health and safety is a pressing concern in the mining industry worldwide. The World Health Organization (WHO) defines health as a state of complete physical, mental, and social well-being (Leonardi, 2018). A healthy workplace recognizes and controls health risks. The Factories Act on Occupational Health and Safety in Zambia addresses these issues, requiring employers to provide a clean working environment with adequate ventilation, first aid, lighting, sanitary facilities, and fire extinguishers (OHSA, 2010).

The mining industry in Zambia is governed by rules and regulations concerning occupational health and safety. Employers must provide safe systems of work (OHSA, 2010). Regulations also provide for safety and health supervision, inspections, accident investigations, and statistics compilation (ILO, 2013). Employees must take reasonable care for their own health and safety and cooperate with the employer to enable compliance (ILO, 1996; Laws of Zambia, 2018).

Accidents and injuries are a leading cause of death in Zambia (Zambwe et al., 2021). Despite regulatory frameworks and OHS services, uncertainties persist regarding their efficacy in minimizing risks and ensuring workers’ welfare. Studies have assessed accidents and injuries in copper mines in Zambia (Malama, 2018), but there is a lack of information on hazards and safety services in artisanal small-scale gold mines. Mercury-dependent gold mining operations expose workers to significant occupational health and safety hazards (Ottenbros et al., 2019). Mercury exposure can cause severe health effects, including respiratory system damage, kidney damage, and neurological symptoms (Afrifa et al., 2019). This study assessed compliance with OHS services within mercury-based gold mines in Mumbwa District, Zambia, to pinpoint deficiencies, strengths, and opportunities for enhancement to bolster worker protection and foster workplace safety.

## Materials and Methods

### Study Design

This study employed a sequential mixed-methods approach, integrating both quantitative and qualitative data collection and analysis techniques to provide a comprehensive assessment of occupational health and safety (OHS) services in mercury-based gold mines in Mumbwa District, Zambia. The quantitative phase involved a cross-sectional survey to collect data on hazard exposure and OHS compliance from miners. The qualitative phase used in-depth interviews to explore the experiences and perspectives of safety supervisors regarding facilitators and barriers to OHS compliance. The data collection process took place in December 2024.

### Study Setting

The study was conducted in the Mumbwa District of Central Province, Zambia. Mumbwa is known for its active artisanal and small-scale gold mining operations, where mercury is commonly used in the gold extraction process. The selection of Mumbwa was based on the prevalence of mercury-based Artisanal small-scale gold mining activities and the associated health risks reported in the area.

### Study Population

The study population consisted of two primary groups: miners and safety supervisors. The miners included individuals directly involved in gold mining activities, particularly those who used mercury in the gold amalgamation process, placing them at potential risk of exposure. The safety supervisors comprised personnel responsible for overseeing mining operations with a focus on implementing and enforcing occupational health and safety (OHS) standards within the gold mines.

### Sample Size and Sampling Technique

A total of 108 miners were involved in the study from the various gold mining sites in Mumbwa District. These respondents were convenience included in the study based on availability, and difficulty in obtaining a comprehensive list of all miners in the area, given the informal nature of many mining operations. For the qualitative phase, purposive sampling was used to select safety supervisors who had extensive knowledge of OHS practices in the mines. A total of 10 safety supervisors were interviewed. This number was arrived at based on data saturation, where no new themes or insights emerged from additional interviews.

### Data Collection Instruments

Data collection for the study utilized a combination of structured questionnaires, in-depth interviews, and observational checklists to gather comprehensive information. Structured questionnaires were administered to miners to collect data on demographics, occupational hazards experienced, awareness of occupational health and safety (OHS) practices, and access to OHS services. Semi-structured, in-depth interviews were conducted with safety supervisors to obtain detailed insights into OHS policies, challenges faced in implementation, and factors influencing compliance. Additionally, an observational checklist was employed to assess the physical conditions of the mining sites, the availability of safety equipment, and the degree of adherence to safety protocols. This mixed-method approach ensured a thorough understanding of both self-reported and observed aspects of occupational health and safety within the gold mining environment.

### Data Collection Procedure

#### Quantitative Data Collection

Obtained informed consent from miners before administering the questionnaire. Administered questionnaires face-to-face to ensure clarity and completeness of responses. Collected data on demographics, occupational hazards, awareness of OHS practices, and access to OHS services.

#### Qualitative Data Collection

Scheduled in-depth interviews with safety supervisors at a convenient time and location. Conducted semi-structured interviews using a pre-defined interview guide. Recorded interviews with permission from the participants. Took detailed notes during the interviews to capture non-verbal cues and contextual information.

#### Observational Data Collection

Conducted site visits to the gold mines accompanied by a trained observer.

Used the observational checklist to assess the presence of safety equipment, adherence to safety protocols, and overall working conditions.

### Data Management and Storage

All datasets were securely stored on a password-protected computers with restricted accessibility. Additional copies were backed up on a secured Google Drive account with restricted access to ensure confidentiality. Antivirus protection was kept up to date to prevent data loss from malware or hardware failure. These measures were taken to maintain the integrity, confidentiality, and retrievability of the data.

### Data Analysis

#### Quantitative Data Analysis

Data from the questionnaires were entered into a statistical software package (e.g., SPSS) for analysis. Descriptive statistics (frequencies, percentages, Odds ratios, and P-values) were used to summarize demographic characteristics, hazard exposure, satisfaction of workers to the OHS services provided, and OHS compliance levels.

#### Qualitative Data Analysis

Interview recordings were transcribed verbatim. Thematic analysis was used to identify recurring themes and patterns in the data. Data were coded and categorized to facilitate theme development. Representative quotes were selected to illustrate key findings.

#### Integration of Quantitative and Qualitative Data

Quantitative and qualitative findings were integrated during the interpretation phase to provide a more nuanced understanding of OHS compliance in the gold mines. Qualitative data were used to contextualize and explain quantitative findings.

### Ethical Approval and Consent to Participate

This study was conducted in accordance with the ethical principles set forth in the World Medical Association Declaration of Helsinki. Prior to commencement, the research protocol was reviewed and approved by the University of Zambia Biomedical Research Ethics Committee (UNZABREC-Approval number 5738-2024; Date 18th September, 2024), and also permission to carry out the research was granted by the National Health Research Authority (NHRA) of Zambia with the registration number NHRA-R-1608/27/05/2024. Written informed consent was obtained from all participants, who were adequately informed about the study’s aims, procedures, potential risks, and benefits. Participant confidentiality and privacy were strictly maintained throughout the study. The study adhered to principles of minimizing harm and ensuring the welfare and rights of all human subjects involved. Any adverse events were promptly managed and reported. The researchers declare adherence to all ethical standards in the conduct, analysis, and reporting of this research.

## Results

### Socio-Demographic Factors

The demographic data of the study participants revealed that out of the total sample, 102 participants, majority of participants fell within the age group of 25 to 34 years, representing nearly half of the sample at 47.1%. The younger demographic (under 25) also comprised a significant portion at 30.4%. The gender distribution indicated a predominantly male workforce, with males accounting for over 93% of the participants, highlighting a significant gender imbalance in this sector.

**Table 1.**
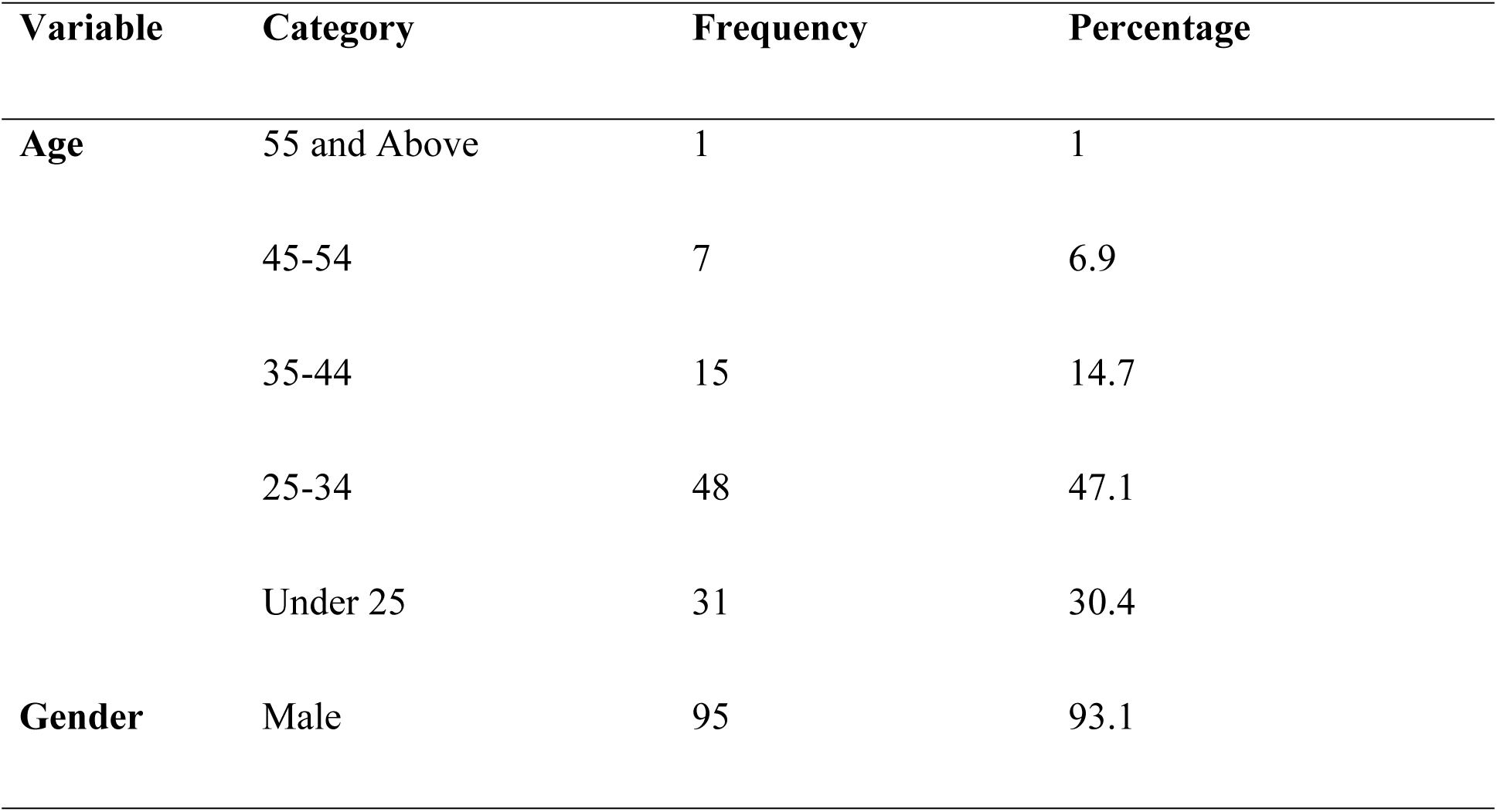
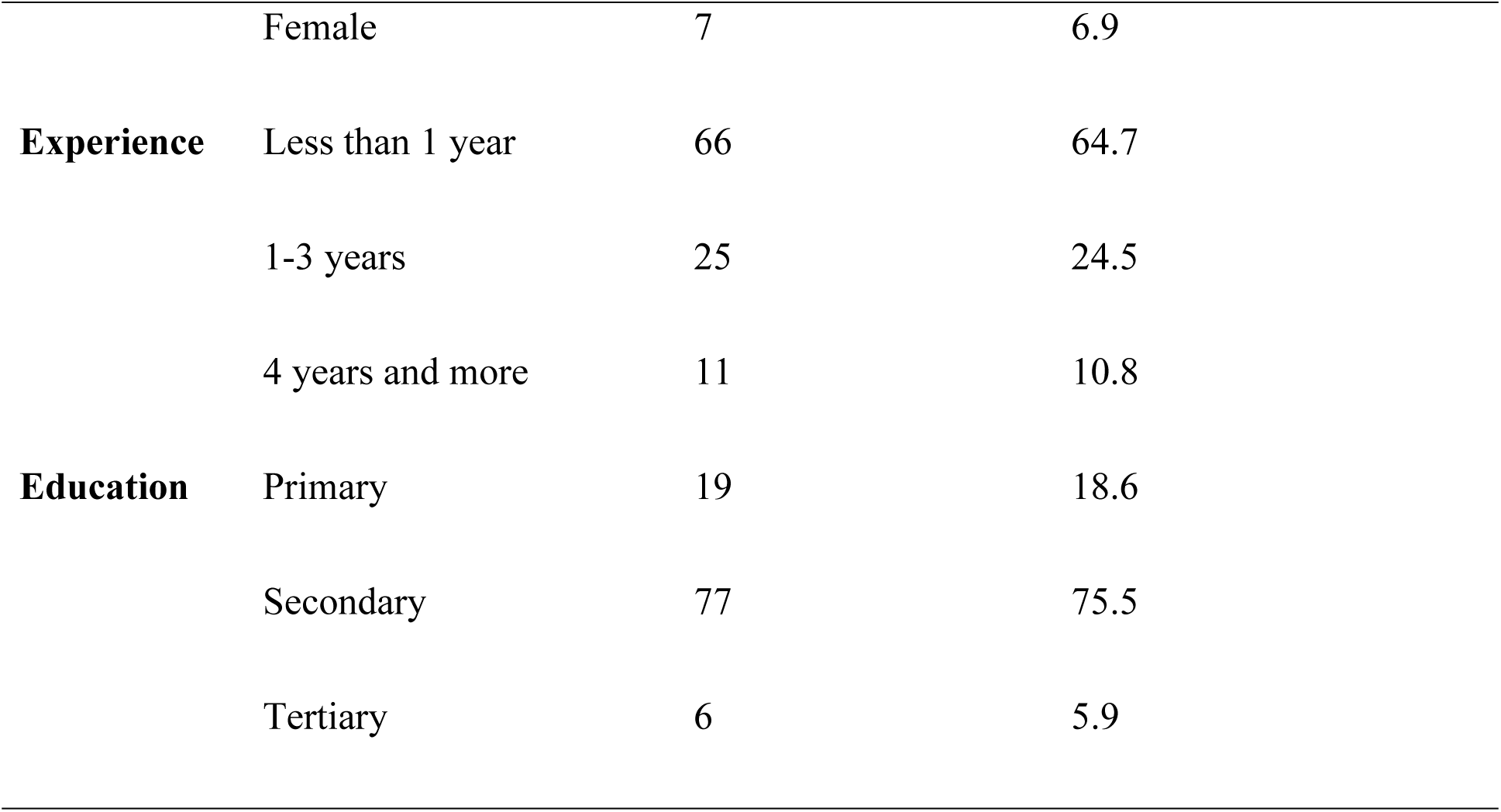
Socio-demographics.

| Variable | Category | Frequency | Percentage |
| --- | --- | --- | --- |
| Age | 55 and Above | 1 | 1 |
|  | 45-54 | 7 | 6.9 |
|  | 35-44 | 15 | 14.7 |
|  | 25-34 | 48 | 47.1 |
|  | Under 25 | 31 | 30.4 |
| Gender | Male | 95 | 93.1 |
|  | Female | 7 | 6.9 |
| **Experience** | Less than 1 year | 66 | 64.7 |
|  | 1-3 years | 25 | 24.5 |
|  | 4 years and more | 11 | 10.8 |
| **Education** | Primary | 19 | 18.6 |
|  | Secondary | 77 | 75.5 |
|  | Tertiary | 6 | 5.9 |

### Hazard Profile

Respondents were asked to state the common occupational health and safety hazards in the gold mines. A significant majority of respondents (98%) reported exposure to dust and particulate matter, while only a small percentage (7.8%) acknowledged noise as a hazard, but majority (92.2%) did not perceive it as a significant risk, which may warrant further investigation into noise levels and their potential impact on workers’ health. Vibration exposure was reported by only 4.9% of respondents, with a substantial majority (95.1%) indicating no exposure. Over half of the participants (56.9%) reported experiencing chemical exposure. Only 14.7% of respondents indicated exposure to heavy machinery hazards, while a large majority (85.3%) reported no such exposure, suggesting effective management practices may be in place. A notable portion (62.7%) reported mercury exposure. Ergonomic hazards were identified by only 9.8% of respondents, suggesting that this may not be a widespread issue. Approximately one-fifth (21.6%) of respondents acknowledged heat stress as a hazard.

**Table 2.** Hazard Profile.

| **Hazard** | **Category** | **Frequency** | **Percentage** |
| --- | --- | --- | --- |
| Dust and Particulate matter | Yes | 100 | 98 |
|  | No | 2 | 2 |
| Noise | Yes | 8 | 7.8 |
|  | No | 94 | 92.2 |
| Vibration | Yes | 5 | 4.9 |
|  | No | 97 | 95.1 |
| Chemical Exposure | Yes | 58 | 56.9 |
|  | No | 44 | 43.1 |
| Heavy Machinery | Yes | 15 | 14.7 |
|  | No | 87 | 85.3 |
| Mercury exposure | Yes | 64 | 62.7 |
|  | No | 38 | 37.3 |
| Ergonomic Hazards | Yes | 10 | 9.8 |
|  | No | 92 | 90.2 |
| Heat Stress | Yes | 22 | 21.6 |
|  | No | 80 | 78.4 |

### Observed Occupational Hazards and Their Profiles

Respondents were asked to state the common occupational health and safety hazards in the gold mines. A significant majority of respondents (98%) reported exposure to dust and particulate matter, while only a small percentage (7.8%) acknowledged noise as a hazard, but majority (92.2%) did not perceive it as a significant risk, which may warrant further investigation into noise levels and their potential impact on workers’ health. Vibration exposure was reported by only 4.9% of respondents, with a substantial majority (95.1%) indicating no exposure. Over half of the participants (56.9%) reported experiencing chemical exposure. Only 14.7% of respondents indicated exposure to heavy machinery hazards, while a large majority (85.3%) reported no such exposure, suggesting effective management practices may be in place. A notable portion (62.7%) reported mercury exposure. Ergonomic hazards were identified by only 9.8% of respondents, suggesting that this may not be a widespread issue. Approximately one-fifth (21.6%) of respondents acknowledged heat stress as a hazard.

**Table 3.**
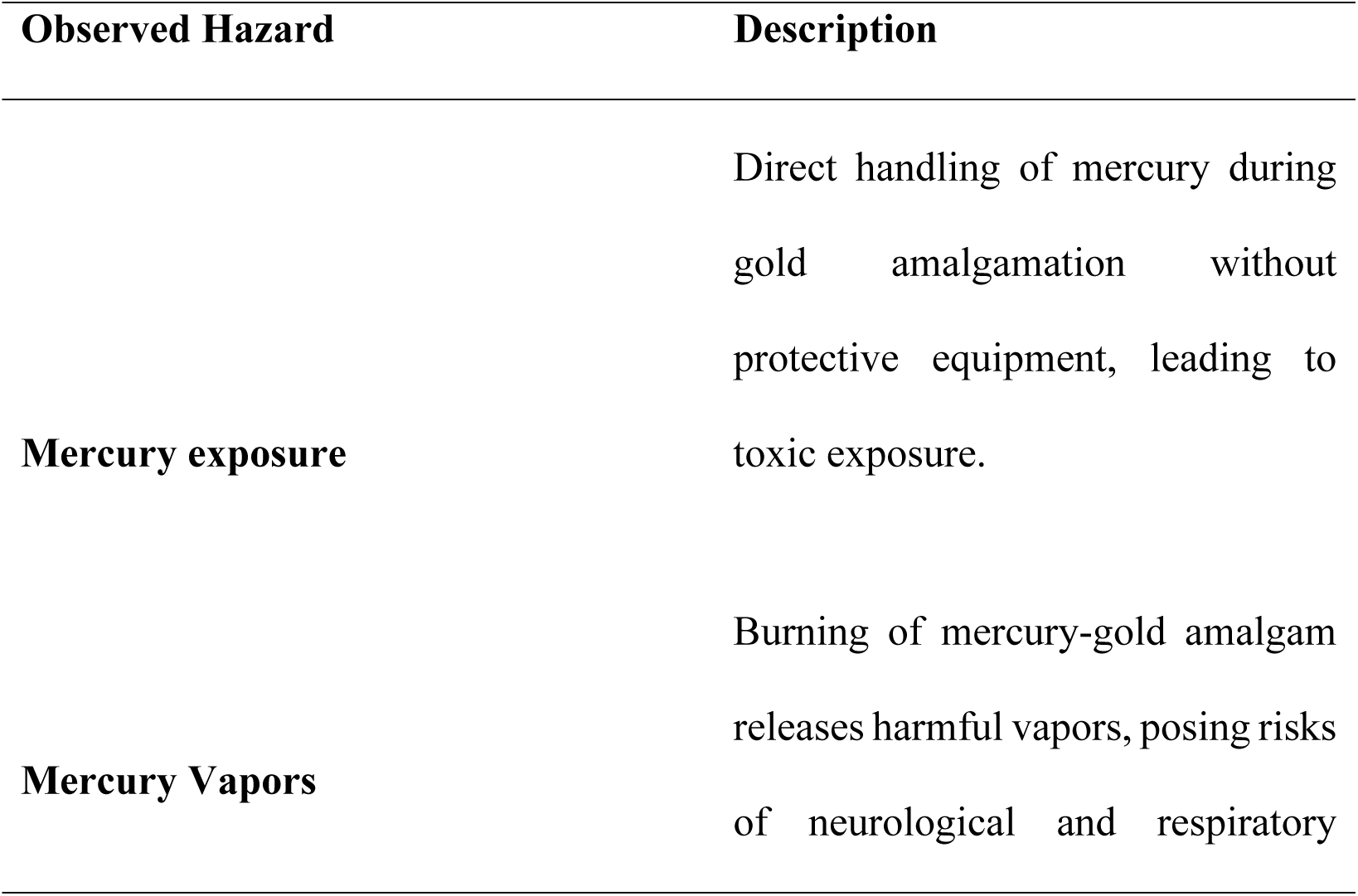
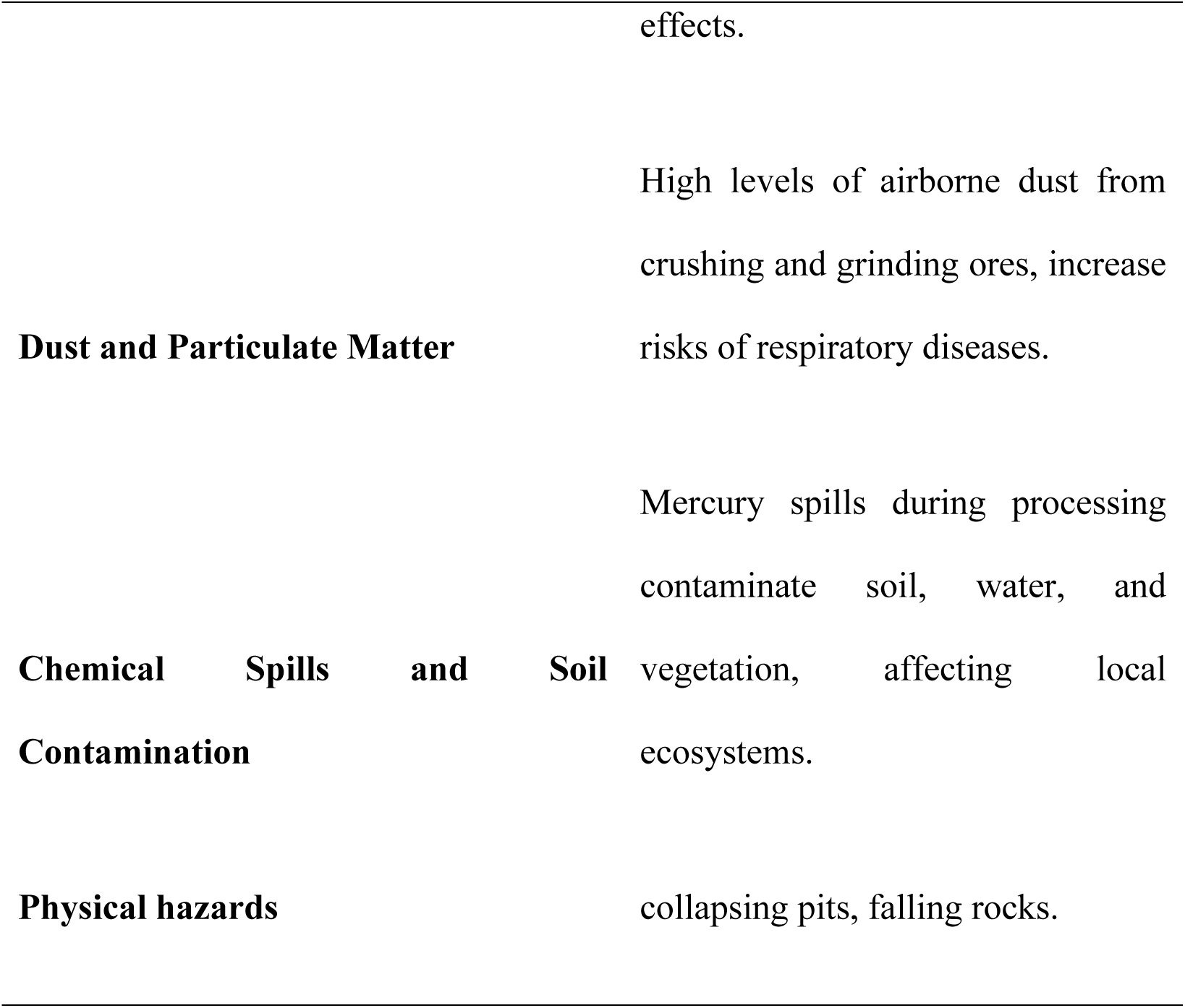
Observed Occupational Hazards and Their Profiles.

| Observed Hazard | Description |
| --- | --- |
| Mercury exposure | Direct handling of mercury during gold amalgamation without protective equipment, leading to toxic exposure. |
| Mercury Vapors | Burning of mercury-gold amalgam releases harmful vapors, posing risks of neurological and respiratory |
|  | effects. |
| **Dust and Particulate Matter** | High levels of airborne dust from crushing and grinding ores, increase risks of respiratory diseases. |
| **Chemical Spills and Soil Contamination** | Mercury spills during processing contaminate soil, water, and vegetation, affecting local ecosystems. |
| **Physical hazards** | collapsing pits, falling rocks. |

### Available Occupational Health and Safety Services in the Artisanal Small-scale Gold mines

Respondents were asked about the presence or the availability of several occupational health and safety services. When asked about training, only 24.5% of respondents reported that training on occupational health and safety was present. A mere 32.4% indicated that regular health checks were available, while 67.6% reported their absence. Regarding onsite medical services, Alarmingly, none of the respondents reported the presence of on-site medical services. The presence of a reporting system was acknowledged by 65.7% of respondents while only 15.7% reported having an OHS safety committee in place.

**Table 4.**
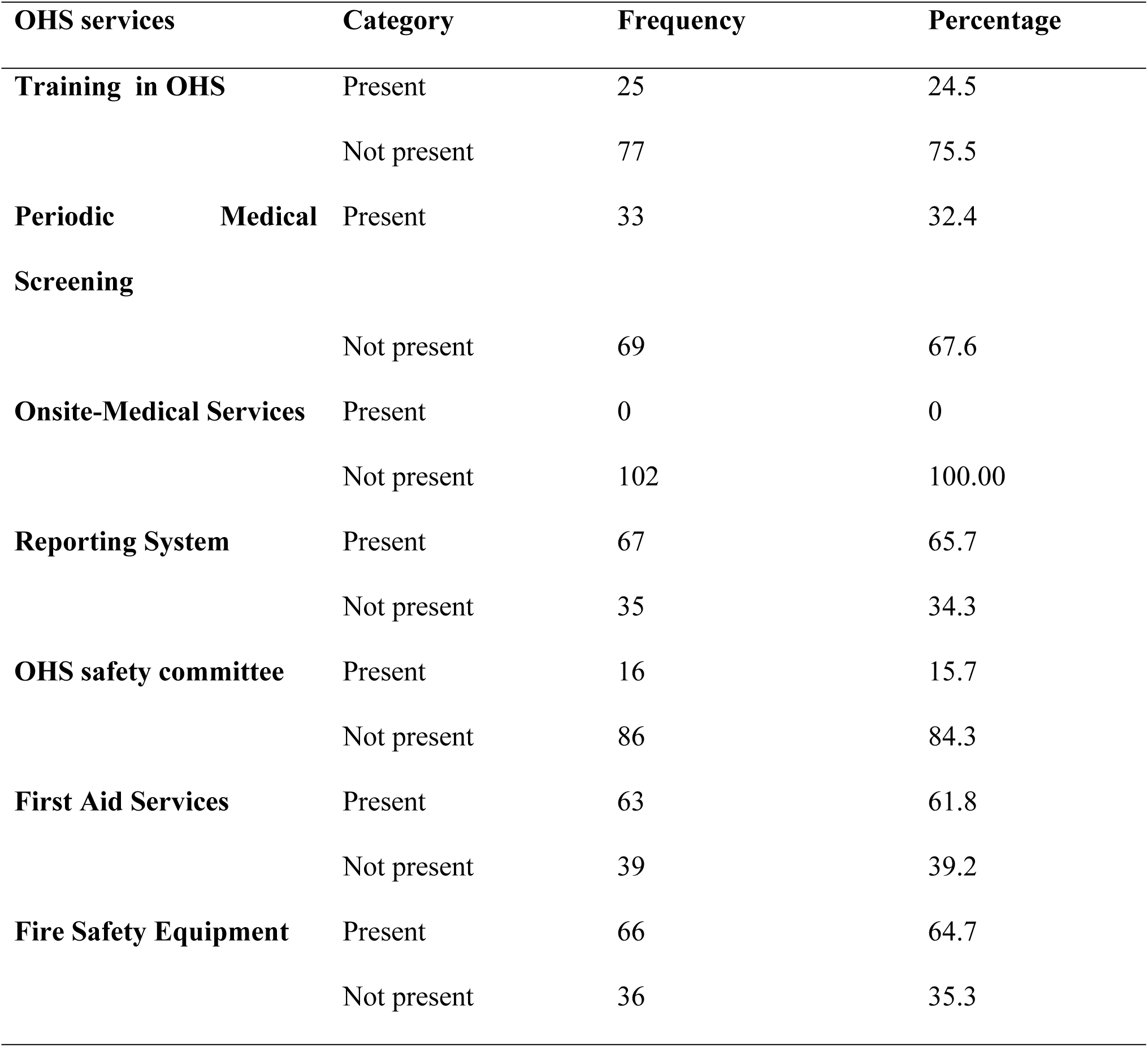
Occupational Health and safety Services in *the Artisanal Small-scale Gold mines*.

### Compliance Rate Across Different OHS Standards by the Mines

The artisanal small scale gold mines were assessed for compliance to the different OHS standards. Only 23.8% of the mines reported compliance with training standards. Compliance with health checks stands at 33.3%, and no mines reported having onsite medical services available. A relatively higher compliance rate of 66.7% was observed for reporting systems. Compliance with the establishment of OHS safety committees is notably low at 14.3%. The compliance rate for first aid services is 62%. Fire safety measures are reported to be compliant in 66.7% of the mines.

**Table 5.**
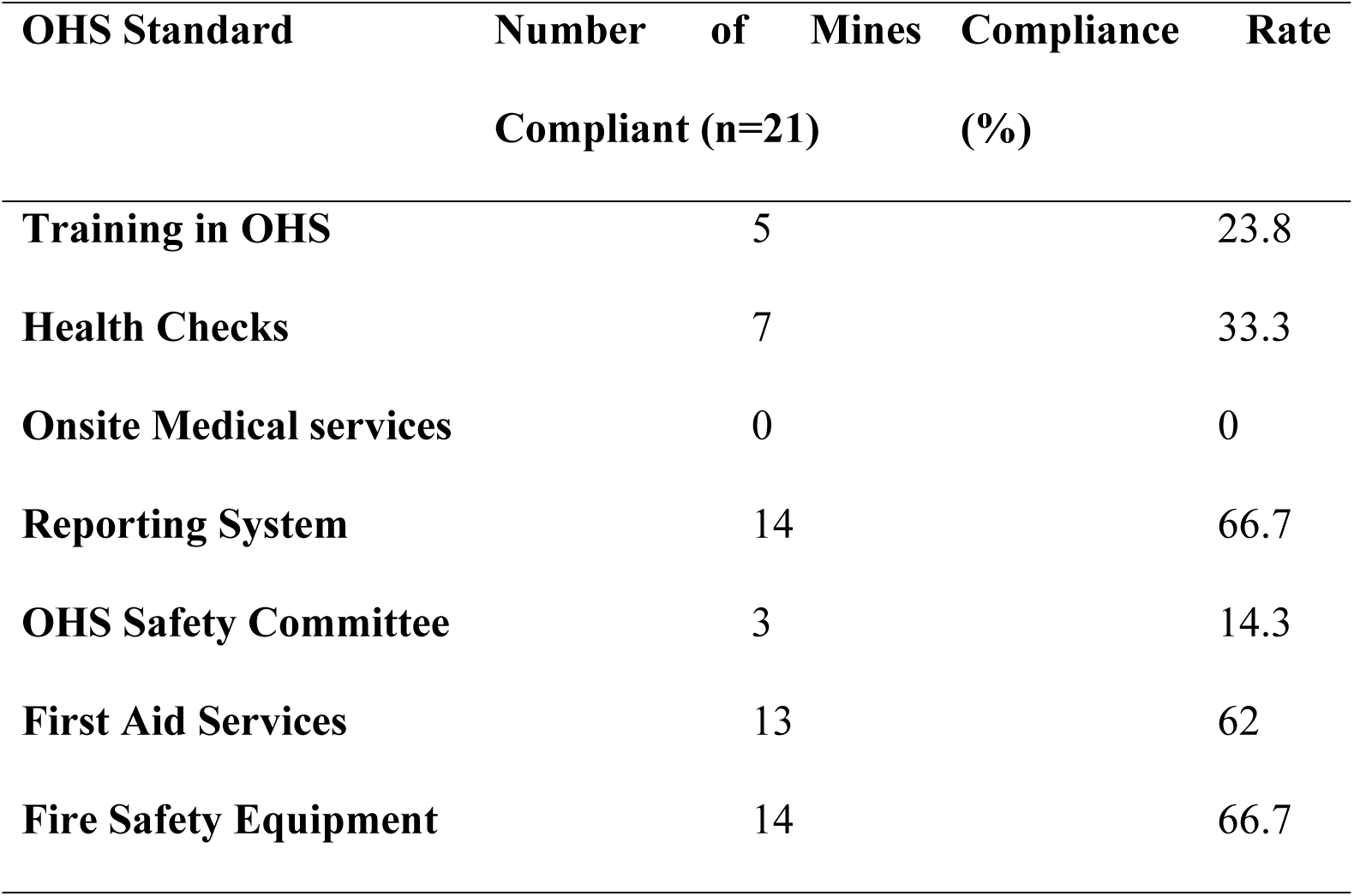
Compliance with OHS Standards by the mines.

### Satisfaction of the Workers to the OHS services in the mines

The analysis identified several factors associated with worker satisfaction at 5% confidence interval. Training emerged as the strongest predictor, with workers receiving training having 4.8 times higher odds of reporting greater satisfaction compared to untrained workers (OR = 4.82, 95% CI: 1.74–13.33, p = 0.002). Similarly, access to First Aid services significantly increased satisfaction odds by nearly 5.9 times (OR = 5.88, 95% CI: 1.66–20.86, p = 0.006). Experience also showed a positive association, with more experienced workers having 2.75 times higher odds of satisfaction (OR = 2.75, 95% CI: 1.07–7.11, p = 0.036). While Education demonstrated a trend toward higher satisfaction (OR = 1.92, 95% CI: 0.89–4.17), its effect was non-significant (p = 0.097). Gender (OR = 2.25, p = 0.240) and Fire safety measures (OR = 0.96, p = 0.944) did not show statistically significant associations, with confidence intervals spanning the null value (1), indicating uncertainty in their effects.

**Table 6.**
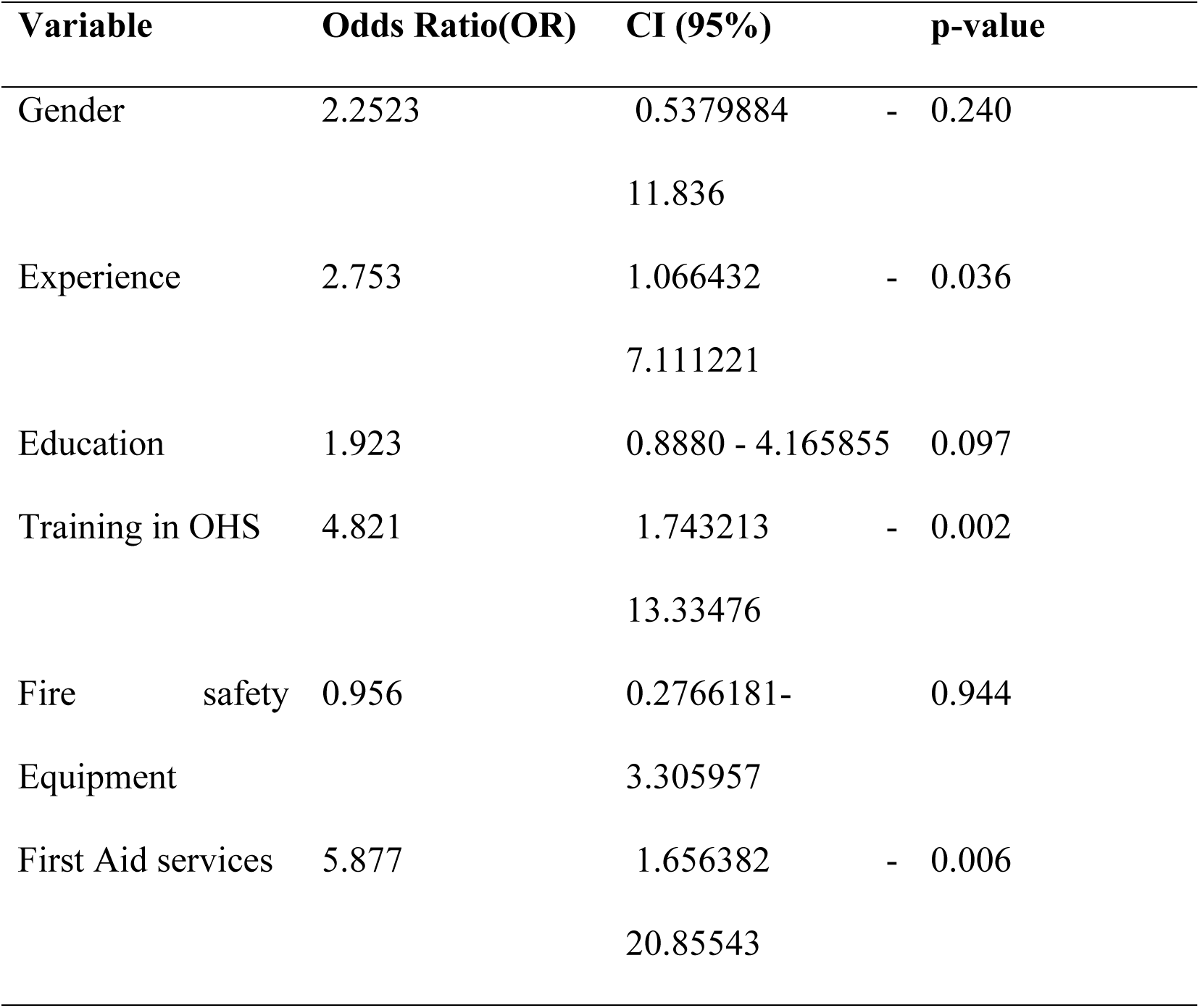
Factors associated with worker’s Satisfaction.

### Facilitators and Hindrances to Compliance with OHS Standards

The Occupational health and safety personnel showed varying views of the factors that facilitate compliance with OHS standards. Some indicated that safe work environments, regulations, enforcement and management support are key aiders to compliance. This is what one had to say on compliance to OHS standards:

> *So there are a lot of factors I would say that would help probably for us to identify whether a company is being compliant or not. And first and foremost, it’s the general environment of the workplace, I would say. So as a safety officer, it’s my responsibility to make sure that someone is working in a safe environment and the work has to be done in a safe way. So in the end, we need to be safe and even the environment needs to be safe and even our surrounding is not supposed to be affected with the activities that we are doing. So we need to take care of the processes, the people themselves, the equipment that we are using. I think those are some of the factors that can help us identify*. – IDI number 2

Another had this to say pertaining to the role that management plays in compliance:

> *Management supporting is also another thing that helps compliance. Like in our company, the management is very supportive when it comes to safety and health compliance. Because at times you find that maybe we need something that will help to facilitate safety and health. For instance, maybe, let’s say we want personal protective equipment like safety boots, reflective vests, gloves, and other things. They respond positively*. - IDI number 1

### Challenges to Compliance with OHS Standards

The occupational health and safety personnel who participated in the study also showed that several factors present challenges to compliance. These include lack of financial resources and resistance to change by the workers. This is what one had to say:

> *So, challenges are many because Occupational Health and Safety, despite it being a good program, it involves financial matters issues. So, where finances are concerned, companies or organizations will definitely have issues. Where is good production, that’s where they put most of it. So others would think they are wasting or they are misusing resources simply because we are trying to invest in Occupational Health and Safety. So, the major thing would be finances because they would rather invest in something else. Then safety becomes secondary*. – IDI number 2

## Discussion

The age distribution revealed that a significant portion of the participants fell within the 25 to 34-year age group. This indicates a youthful workforce, which could imply a need for targeted health and safety training and awareness programs tailored to younger miners. In contrast, only a small percentage of the respondents were aged 55 and above, suggesting that older workers are less represented in these mining operations. Since, most of the population are youthful, chronic mercury exposure in artisanal gold mining (ASGM) may pose severe, long-term health risks due to their developing physiology and heightened vulnerability. Mercury damages retinal and auditory nerves, leading to blurred vision or tinnitus later in years. The results reveal that most of the respondents fall within the working age. Thus within 18-35 and 36-50. This is in line with the findings of (Siabi et al., 2023) which revealed that ASGM is highly dominated by the youth. This may be attributed to the physical and strenuous nature of the ASGM activities which demands physical strength. Therefore, compared to large-scale mining, ASGM is less capital-intensive but demands more manpower. The gender distribution highlighted a predominantly male workforce. This significant gender imbalance points to potential barriers for women in accessing employment opportunities within the gold mining sector, reflecting broader trends often seen in artisanal and small-scale mining environments.

In terms of experience, a substantial majority of participants had less than one year of experience in gold mining. This suggests that many workers may be new to the industry, which could impact their understanding of occupational health and safety practices. This also indicates a relatively transient workforce that may require ongoing training and support. The educational background of the participants showed that most had completed secondary education, while only a few had attained tertiary education. This educational profile suggests that while many workers have basic literacy and numeracy skills, there may be a gap in advanced training or specialized education related to occupational health and safety practices. This agrees with the findings of (Arthur et al., 2016). Arthur et al. (2016) noted that since ASGM demands people with less special skills, most of its workers are those in rural areas. Irrelevant employable skills leading to unavailability of other livelihoods than ASGM may be the main cause. The educational levels also suggest an opportunity for further training initiatives to enhance knowledge about occupational health risks associated with mercury use in mining operations.

A significant majority of respondents reported exposure to dust and particulate matter, indicating a serious concern for respiratory health and overall worker safety. The overwhelming acknowledgment of dust exposure underscores the need for effective dust control measures in mining operations. Studies have consistently shown that prolonged exposure to silica dust can result in severe respiratory diseases, emphasizing the importance of implementing adequate ventilation systems and dust suppression techniques (Verma et al., 2014). Over half of the participants reported experiencing chemical exposure, raising significant concerns about the adequacy of safety measures and protective equipment available to mitigate these risks. The International Labour Organization (ILO) has long recognized that workers in artisanal and small-scale gold mining are particularly vulnerable to chemical hazards, including mercury, which can cause severe neurological and physiological effects. Mercury is known to disrupt thyroid function, delaying puberty and stunting growth. Oxidative stress may damage reproductive organs, increasing infertility risks in adulthood. A notable portion reported mercury exposure, indicating a significant occupational health risk that requires immediate attention. Research has shown that mercury exposure can lead to serious health issues, including neurological disorders and cognitive dysfunction (Taux et al., 2022). The findings underscore the urgent need for implementing protective measures such as proper ventilation, personal protective equipment (PPE), and training on safe mercury handling practices.

Majority of the mines were moderately compliant to the occupational health and safety standards with respect to the seven parameters examined in this study. These include fire safety, first aid services, reporting system, OHS safety committee, onsite medical services, health checks and training. The low compliance rate for training highlights a significant gap in educating workers about safety practices, which are essential for reducing accidents and improving safety culture. The limited availability of health checks reflects insufficient monitoring of workers’ health, particularly concerning mercury exposure. Mercury is known to cause severe neurological, respiratory, and systemic health effects, even at low levels of chronic exposure. The complete absence of onsite medical services poses a serious risk to miners, as immediate medical intervention is critical in cases of acute mercury poisoning or mining-related injuries. This mirrors findings from the Mine Safety and Health Administration (MSHA), which notes that many small-scale mining operations globally lack basic healthcare infrastructure to address emergencies. The lack of onsite medical services undermines the ability to manage acute health crises effectively, leaving miners vulnerable to preventable morbidity and mortality. The relatively higher compliance rate for reporting systems indicates some progress in hazard identification and incident reporting mechanisms. Effective reporting systems are vital for identifying risks and implementing corrective actions. The low compliance rate for establishing OHS safety committees reflects a lack of worker participation in promoting workplace safety. Safety committees are crucial for fostering communication between management and employees regarding health and safety concerns. According to Grossman (2018), poor communication can lead to employee mistrust, absenteeism and low morale, bad interpersonal relationships, and the grapevine effect. On the contrary, good communication can help to achieve productivity and maintain strong working relationships at all levels in the mines. If the mines invested time and energy into delivering clear lines of communication, they would rapidly build trust among the employees, leading to increases in productivity, output and morale in general (‘Communication Barriers in Work Environment: Understanding Impact and Challenges’, 2024). Gaps in compliance with occupational health and safety standards can lead to increased workplace injuries, illnesses, and fatalities by exposing workers to preventable hazards. Furthermore, such non-compliance undermines employee morale and productivity, while also increasing legal liabilities and financial costs for employers.

The qualitative results from the study on compliance with occupational health and safety (OHS) standards in mercury-based gold mines in Mumbwa district reveal both facilitators and challenges that significantly influence adherence to these standards. The study indicated that a safe work environment is crucial for compliance. This aligns with findings from other studies that underscore the importance of workplace safety culture in enhancing compliance with OHS standards (Cohen et al., 2018). The presence of government regulations and enforcement mechanisms was identified as a facilitator of compliance. One participant mentioned that “regulations…help us to enforce” compliance. This reflects broader findings in occupational health literature, which suggest that effective regulatory frameworks are essential for ensuring adherence to safety standards (López et al., 2019). Regular inspections by enforcement agencies were highlighted as beneficial for identifying and rectifying compliance issues promptly. Respondents noted that these inspections carry significant weight and encourage timely corrective actions by management. This supports previous research indicating that regular audits and inspections can lead to improved safety practices.

The findings underscored the critical role of targeted interventions—particularly training and access to First Aid—in improving worker satisfaction in mercury-based gold mines. Experience also contributes positively, while education, gender, and fire safety measures did not show statistically significant effects in this analysis. These insights can inform policy and management strategies aimed at enhancing worker well-being in hazardous mining environments.

A significant barrier to compliance identified in the study was the lack of financial resources. Respondents indicated that companies often prioritize production over safety investments, viewing expenditures on OHS as secondary or unnecessary. This finding resonates with other studies that have reported financial limitations as a common obstacle to implementing effective OHS measures (Krause et al., 2018). These results were consistent with the findings of a study on costs of compliance in relation to the benefits of compliance amongst Small and Medium-sized Enterprises (SME’s) by Chan et al., (2016) which similarly found out that regulatory compliance is usually very low when the costs of complying with rules in terms of time, money and effort are considered to be high. In another study similar study by Kalidin, (2017) the cost of compliance was found to be embedded in the complex nature of rules and the general regulatory structures. Complex nature of rules and general regulatory structure essentially raise compliance cost in relation to turnover consequently affecting compliance rates. Lastly, this study concurred with the findings of a study by (Lyon & Maxwell, 2016) from majority of the OECD countries which concluded that the cost of compliance is usually higher for SME’s, thus are at a higher risk of experiencing compliance failure.

## Conclusion

This study highlights significant gaps in OHS compliance in mercury-based gold mines in Mumbwa District, Zambia. Miners face high levels of exposure to dust, chemicals, and mercury, while access to adequate training, health checks, and medical services is limited. Addressing these deficiencies requires a concerted effort involving regulatory bodies, mine owners, and miners themselves. Implementing comprehensive OHS strategies, strengthening enforcement mechanisms, and promoting a culture of safety are essential to protect the health and well-being of miners in Mumbwa District and beyond. Future research should focus on evaluating the effectiveness of specific OHS interventions and exploring sustainable solutions to improve working conditions in artisanal gold mines.

## Data Availability

The data that support the findings of this study are available from the corresponding author Musole Philip upon reasonable request.

## Supporting Information

The research work possesses no additional information aside from what has been communicated.

## Acknowledgements

The authors thankfully acknowledge the University of Zambia, School of Public Health, Department of Environmental Health, and the Zambia Environmental Management Agency (ZEMA) for their time, support, and Collaboration.

